# The role of Remote Clinical Care in Health Promotion: a scoping review protocol

**DOI:** 10.1101/2025.10.01.25337066

**Authors:** Craig Brown, Nathan Bray, Mike Brady, Lorelei Jones

**Affiliations:** Welsh Ambulance Services University Trust. St Asaph, Denbighshire, Wales, United Kingdom; College of Medicine and Health, Bangor University, Wales, United Kingdom

**Author notes:** Corresponding Author: Craig Brown.

**Keywords:** Remote Care, Telephone Triage, Telehealth, Digital Healthcare, Virtual Care, Health Promotion, Health Education, NHS 111

## Abstract

**Objective:** This scoping review aims to understand what opportunities exist to provide health promotion to the adult population within remote clinical care.

**Introduction:** With increasing demand for healthcare in an environment of overstretched healthcare resources, and the legal responsibility on public organisations within Wales to improve long-term health, all opportunities to promote health should be considered. The consolidation of remote clinical care in most western healthcare services may provide such potential opportunities.

**Inclusion criteria:** The inclusion criteria will incorporate adults contacting remote clinical care services that provide health promotion. It will exclude any sources not written in English, restrict articles to post-2013, and exclude articles that focus entirely on mental well-being or individuals under 16.

**Methods:** Searches will be undertaken in Medline (PubMed), CINAHL (EBSCOhost), Embase, Web of Science, Scopus and British Nursing databases to identify published studies. Searches will also be undertaken on Google Scholar, Google, ProQuest Dissertations, and Theses Global, as well as direct contact with NHS 111 (or equivalent) service providers within the UK to identify any grey literature. Searches will also include bibliographic searches of published studies.

The JBI methodology for Scoping Reviews will be followed, and data extraction, analysis, and presentation of the results will follow the laid-out principles. As this review forms part of a wider Doctorate in Public Health submission, data screening and selection will be undertaken by the primary author only, and supervision for the doctorate will be undertaken by the other three authors.

**Review registration number:** Open Science Framework: osf.io/gqh83

## Introduction

Good health and well-being result in positive outcomes for the individual, their family, community and wider society ^(1)^. The United Kingdom, and Wales specifically, like many countries, are increasingly turning their focus to health inequalities and health promotion ^(2)^ ^(3)^ ^(4)^ ^(5)^. The approach is not new ^(6)^ ^(7)^ ^(8)^ ^(9)^. In 1998, the World Health Organisation (WHO) defined health promotion as the process of enabling people to increase control over, and to improve, their health. Whilst this definition has not formally changed, it has been developed into principles and actions for health promotion. More recently, the Shanghai declaration (2016) provided a transformational framework utilising health promotion to help achieve the UN Sustainable Development Goals ^(10)^.

Viner and Macfarlane ^(11)^ *have long defined health promotion as* ‘the science or art of helping people change their lifestyle to move towards a state of optimal health. Lifestyle change can be facilitated through a combination of efforts to increase awareness, change behaviour, and create environments that support good health practices.’ Prevention is considered more important than ever due to the demographic challenges that increase pressure on health and care services within Wales ^(2)^.

### Welsh demographics

Wales is a part of the United Kingdom, with a population of 3.1 million, 51.1 % female and 48.9% male, and 21.3% are over 65. Population density is relatively low, with a mix between high levels of concentration in the south, e.g., Cardiff, Swansea, Newport, and much more rural areas, specifically in mid Wales ^(12)^. Due to the varying environments, this makes healthcare provision a challenge, presenting a mixed picture of health inequalities and their causes.

### Health promotion in Wales

In the February 2025 regular public engagement survey by Public Health Wales, ‘Time to Talk Public Health’ ^(13)^, involving 2,137 respondents 16 years and above, 80% of respondents reported that they believed primary care services have a role to play in supporting the population to experience good health and to help reduce health inequalities. Furthermore, 85% of respondents stated they would be happy to be asked about their overall well-being, their state of being comfortable, healthy and happy, and 81% believed that healthcare professionals working in primary care environments should have a responsibility for helping patients to eat a healthy diet and be physically active. In the context of the NHS, primary care can be defined as a patient’s first point of contact and secondary care as planned, urgent or emergency care usually based around a hospital setting. Many of the remote care services provided by NHS 111 Wales fall within these definitions ^(14)^.

The NHS Wales Planning Framework ^(2)^ emphasises the need to meet these challenges with greater access to services. Wales will also become a Marmot Nation ^(15)^, focusing more on reducing health inequalities. Good health is not solely the responsibility of the NHS, and includes fundamental foundations based in financial rewards, education, safe and warm housing, and environmental conditions. Harmful levels of alcohol consumption, smoking, poor diet, and inadequate levels of physical activity are key health risk factors, driving high levels of premature mortality from preventable diseases ^(16)^. A Healthier Wales ^(17)^, the Welsh Government’s long-term vision for a system-wide health and social care approach to wellbeing and health prevention, outlined the principles for the population of Wales in managing their own health and preventing illness, independently supported by new technologies and by an integrated health and social care system.

Smoking is the leading cause of preventable ill health and premature death in Wales, with 13% of the Welsh population reported as being smokers, and whilst the rationales for smoking are complex, there are higher rates of smokers living in deprived communities ^(18)^. 61% of adults in Wales are overweight, and 24% are defined as obese ^(19)^. More critically, as obesity increases with age and becomes increasingly difficult to reverse, Welsh children have the highest levels of obesity in the UK, approximately 25% can be considered overweight by school starting age. The National Survey for Wales 2022-23 Sport and Active Lifestyles: State of the Nation Report reported that less than 40% of adults participated in a sport and/or physical activity three or more times per week in Wales, as per the Wellbeing of Future Generations (Wales) Act 2015, National Indicator (No. 38) ^(20)^.

### Digital Health

Remote care has foundations in many clinical fields and specialities, employs a wide range of technologies, and is referred to by a wide range of terminology, including telemedicine, telephone triage, video consultations, and digital healthcare. Across the UK, it emerged mainly from NHS Direct in 1998 and, later, NHS 111 services, driven by the increasing demands and pressures on GP and emergency department services ^(21)^. NHS 111 seeks to provide advice and signposting services to patients 24/7 using non-registered call advisors and clinicians, mainly nurses and paramedics, supported by specialist Clinical Decision Software Systems (CDSS) ^(22)^. However, remote care is increasingly being used in many healthcare arenas, specifically driven during the Covid-19 pandemic as a method to provide care whilst maintaining segregation due to infectious reasons; however, it is more commonly seen as a more efficient and effective method of delivering many forms of care ^(23)^ ^(24)^.

### A scoping review proposal

In July 2025, using the search strategy in Appendix 1, a search of Medline (OVID), PROSPERO, the Cochrane Database of Systematic Reviews, and the JBI Evidence Synthesis was performed, and no complete or in-progress reviews of a similar nature were identified.

The scoping review methodology is typically used to review published studies, evidence and grey literature, including third sector publications, health organisation data and government policies. Due to the review question, the nature of the review, and the varied and assorted documents to be investigated, a scoping review has been identified as an appropriate methodology for this project ^(25)^. This scoping review will identify and describe gaps in the literature, map any potential evidence, inform the discussion on how remote care supports the advancement of health promotion, what strategies have been adopted in this field to improve the delivery and impact of health promotion, and the approaches and theories that have been utilised to provide a foundation for any evaluations.

The scoping review will be undertaken per JBI guidelines for scoping reviews ^(25)^. The review will focus on high-income countries ^(26)^ to ensure that any research recommendations are focused on the defined context of Remote Care in Wales, and the nature of disease patterns experienced by these populations, as this will increase the applicability of the review to similar countries. The included literature will be limited to documents written in English only. It will limit the date range to post 2013, as this marked the rollout of the NHS 111 services in the UK, and the emergence of structured remote triage services, and consequently ensures relevance to the current service models. The principal author intends to use the scoping review results to inform future research activities as part of the lead author’s Professional Doctorate programme.

#### Review questions

1. What is the role of remote care in providing health promotion?
2. Are there opportunities within remote care to provide health promotion?

#### Inclusion criteria

As detailed below, the inclusion criteria were developed using the ‘Participants, Concept, Context’ (PCC) framework.

### Participants

This review will consider published studies and grey literature, including adults who contact remote care services for assessment and advice for any physical health conditions, disease, or injury. Mental health patients will be excluded, as they currently follow different clinical pathways within NHS 111. It will include published studies and grey literature where advice is provided by any healthcare professional; non-registrant healthcare workers will be excluded, as they do not hold a professional registrant’s responsibility to contribute to health promotion. It will also exclude patients previously enrolled in treatment programs, e.g., cardiac rehab, virtual wards.

### Concept

This scoping review will include published studies and grey literature that focus on providing physical health promotion advice within the remote care environment, including telehealth, telephone triage, remote patient monitoring, video health, and digital health. Published studies and grey literature will be included if they provide physical health promotion advice within a remote clinical care environment. Health promotion, as defined by the World Health Organisation ^(10)^, enables people to increase control over and improve their health. In this review of remote care, health promotion can be regarded as aiding patients in modifying their lifestyles to achieve improved physical health. This change can be facilitated by increased awareness, behavioural change and creating environments that support improved health activities ^(11)^. All formal prescriptive, structured as part of the clinical assessment, or informal opportunistic health promotion advice will be reviewed as relevant.

### Context

The remote setting refers to any patient consultation, triage or assessment where the patient and the healthcare professional are not co-located but are within the same country. Remote clinical care can take many forms, including telephony, video, and digital chat assistants. Healthcare professionals may be situated in health facilities, contact centres, or domestic environments. Studies will also be excluded where patients are not within the same country as the healthcare professional service, i.e. aviation medicine.

#### Types of sources

This scoping review will consider all published studies and grey literature, provided they are relevant to the research questions and criteria. These will include experimental and quasi-experimental study designs, including randomised controlled trials, non-randomised controlled trials, before and after studies and interrupted time-series studies. In addition, analytical observational studies, including prospective and retrospective cohort studies, case-control studies and analytical cross-sectional studies, will be considered for inclusion. This review will also consider descriptive observational study designs, including case series, individual case reports and descriptive cross-sectional studies for inclusion.

Qualitative studies, including, but not limited to, designs such as phenomenology, grounded theory, ethnography, qualitative description, and action research, will also be considered for inclusion in this scoping review. Text, opinion papers, conference presentations, and posters will also be considered. It is anticipated that much of the data may come from grey literature, including protocols and standard operating policies from within the UK-wide providers of GP out-of-hours or 111 services.

## Methods

The proposed review has been developed per the JBI methodology for scoping reviews. It will be reported according to the Preferred Reporting Items for Systematic Reviews and Meta-Analysis extension Scoping Reviews (PRISMA-ScR) ^(27)^.

The review has been registered with the Open Science Framework - osf.io/gqh83

### Search strategy

The search strategy will aim to locate both published and unpublished studies. This review will use a three-step search strategy ^(25)^. A preliminary search of Medline (OVID), PROSPERO, Cochrane Database of Systematic Reviews and the JBI Evidence Synthesis was performed in July 2025, and no complete or in-progress reviews of a similar nature were identified. This initial search was undertaken to identify relevant keywords in titles, abstracts, and index terms to develop a complete search strategy (see Appendix 1). The search strategy, including all identified keywords and index terms, will be adapted for each database. The reference list of all included studies will be screened for any additional relevant papers. Due to the context of this review, all documents will be limited to the English language, and a time limit of post-2013 will be set, as it was around this time that the NHS 111 began replacing NHS Direct in England and subsequently Wales ^(24)^.

The databases to be searched include Medline (PubMed), CINAHL (EBSCOhost), Embase, Web of Science, Scopus, and British Nursing. As the search strategy aims to identify grey literature and policy documents, the search will also include ProQuest dissertations and Theses Global, as well as government and health websites. Google and Google Scholar will be searched to identify any other available grey literature. Where data is not publicly available, relevant health services will be contacted for potential resources. Likewise, authors of public literature may be contacted for further information where required.

### Study/Source of evidence selection

Following the search, all identified sources will be collated and uploaded into Mendeley Reference Manager, and any duplicates will be removed. Due to the academic nature of this study, the primary author will screen titles and abstracts of all collected resources to determine whether they are relevant and meet the inclusion criteria. The full text will be read for clarification when there are questions about the resource. The full text of all selected resources will be reviewed against the inclusion criteria, and any full-text articles that are excluded after the review will be recorded and reported in the final review. The search results and the study inclusion process will be reported in full in the final scoping review and presented in a PRISMA-ScR flow diagram ^(27)^. This process will be supported and reviewed by the team of academic supervisors.

### Data extraction

Due to the academic nature of this work, the primary author will extract the data using the JBI Data Extraction Tool ^(25)^. The extracted data will include the authors (s), date reviewed, article type, type of publication, database searched, publication year, study design description, delivery types, health outcomes, participants, interventions, key findings, and the reviewer’s conclusion. Any missing data or ambiguous information will be resolved by contacting the authors or organisations.

### Data analysis and presentation

Relevant data will be extracted from all sources, analysed and published in completed data extraction tool tables, using charting methods to meet the objectives of this scoping review ^(28)^. Descriptive frequencies, counts, and content analysis will be included along with the tabular and graphical results and a narrative summary to describe how the results relate to the objective of this review. Any gaps in the evidence will be highlighted in the findings of this review, which will be used to inform future research projects.

## Data Availability

All data produced in the present work are contained in the manuscript

## Acknowledgements

The principal author (CB) works within the Welsh NHS and is employed in the Welsh Ambulance Service University Trust as a Clinical Development Lead Clinician with a speciality in remote care. The author is undertaking a collaborative Professional Doctorate in Public Health with Bangor University as part of their employment. This review will contribute to the study programme. Other authors (NB, MB, LJ) are the Academic supervisors for the study.

## Funding

Bangor University sponsors the study and provides funding for the Professional Doctorate programme. The Welsh Ambulance Services University Trust funds the study time to undertake the programme. Academic Supervisors represent both the sponsor and funding bodies.

## Declarations

Nil

## Author contributions

CB authored this paper and designed and tested the search strategy. MB, NB, and LJ supervise the Professional Doctorate programme and contributed to the editing and submission of this protocol.

## Conflicts of interest

Nil

## Appendices

## Appendix I: Search strategy

In July 2025, a test search strategy was employed using Medline (Ovid) using the following search terms: The search was limited to English-language documents only, post-2013, adults only, and focused on physical health promotion, not mental health or mental well-being.

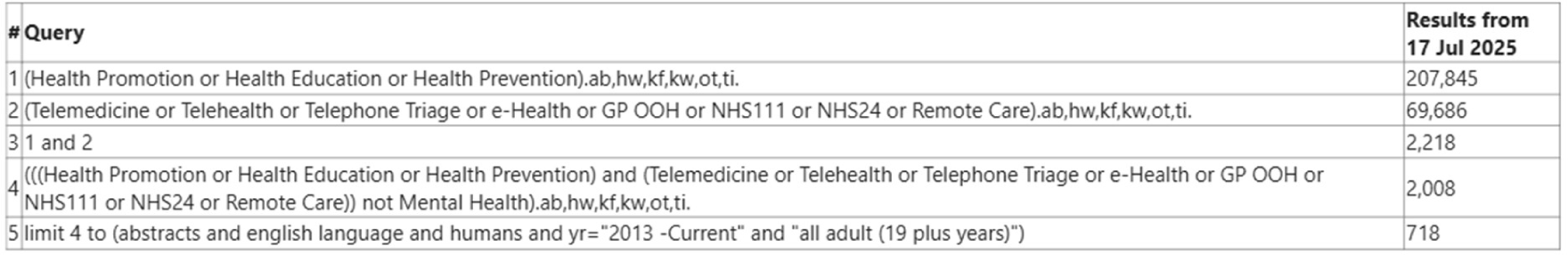

All 718 articles were reviewed using the title and, where required, the abstract. No articles were considered for inclusion in the discussion above.

## Appendix II: Data extraction instrument

The standard JBI data extraction tool will be used within the study.

## Notes

### Competing Interest Statement

The authors have declared no competing interest.

### Funding Statement

This study did not receive any funding

## References

1. Public Health Wales. Investing in a Healthier Wales: prioritising prevention. [internet]. 2025 [cited 2025 August 1]. Available from: https://phwwhocc.co.uk/wp-content/uploads/2025/01/Investing-in-a-Healthier-Wales-prioritising-prevention.pdf

2. Welsh Government. NHS Wales Planning Framework 2022-2025 [internet]. 2021[cited 2025 August 1]. Available from: https://www.gov.wales/sites/default/files/publications/2021-11/nhs-wales-planning-framework-2022-2025_0.pdf

3. Welsh Government. Well-being of Future Generations (Wales) Act 2015: the essentials [internet]. 2025 [cited 2025 August 1]. Available from: https://www.gov.wales/sites/default/files/pdf-versions/2025/2/2/1740501518/well-being-future-generations-act-essentials.pdf

4. UK Government. Fit for the Future: 10-year health plan for England [internet]. 2025 [cited 2025 August 1]. Available from: https://assets.publishing.service.gov.uk/media/6888a0b1a11f859994409147/fit-for-the-future-10-year-health-plan-for-england.pdf

5. Scottish Government. Scotland’s Population Health Framework 2025-2035 [internet]. 2025 [cited 2025 August 1]. Available from: https://www.gov.scot/binaries/content/documents/govscot/publications/strategy-plan/2025/06/scotlands-population-health-framework/documents/scotlands-population-health-framework-2025-2035/scotlands-population-health-framework-2025-2035/govscot%3Adocument/scotlands-population-health-framework-2025-2035.pdf

6. Townsend P, Davidson N, & Whitehead M. Inequalities in Health. The Black Report and Health Divide. Penguin. [1992]

7. Marmot M, Allen J, Boyce T, Goldblatt P, Morrison J. (2020). Health equity in England: The Marmot Review 10 years on [internet]. Institute of Health Equity. 2020 [cited 2025 August 1]. Available from: https://www.instituteofhealthequity.org/resources-reports/marmot-review-10-years-on

8. UK Government. Saving Lives: Our Healthier Nation [internet]. 1999 [cited 2025 August 1]. Available from: https://assets.publishing.service.gov.uk/media/5a7b8c8240f0b62826a044aa/4386.pdf

9. Dahlgren G, Whitehead M. The Dahlgren-Whitehead model of health determinants: 30 years on and still chasing rainbows. Public Health 2021; 199: 20–24.

10. WHO. Health Promotion [internet]. 2025 [cited 2025 August 1]. Available from: https://www.who.int/health-topics/health-promotion#tab=tab_1

11. Vinar R, Macfarlane A. ABC of adolescence: Health promotion. BMJ 2005; 330;527–529.

12. Office for National Statistics. Population and household estimates, Wales: census 2021 [internet]. 2021 [cited 2025 August 1]. Available from; https://www.ons.gov.uk/peoplepopulationandcommunity/populationandmigration/populationestimates/bulletins/populationandhouseholdestimateswales/census2021

13. Public Health Wales. Time to talk Public Health: February 2025 Survey Findings [internet]. 2025 [cited 2025 August 1]. Available from: https://phw.nhs.wales/topics/time-to-talk-public-health/time-to-talk-public-health-panel-publications/publications/time-to-talk-public-health-february-2025-survey-results/

14. NHS England. The Healthcare Ecosystem [internet]. 2021 [cited 2025 August 1]. Available from; https://digital.nhs.uk/developer/guides-and-documentation/introduction-to-healthcare-technology/the-healthcare-ecosystem

15. Welsh Government. Wales to become world’s first ‘Marmott nation’ to tackle health inequalities [internet]. 2025 [cited 2025 August1]. Available from: https://www.gov.wales/wales-become-worlds-first-marmot-nation-tackle-health-inequalities

16. Everest G, Marshall L, Fraser C, Briggs A. Addressing the leading risk factors for ill health: A review of government policies tackling smoking, poor diet, physical inactivity and harmful alcohol use in England [internet]. The Health Foundation. 2022 [cited 2025 August 1]. Available from: 10.37829/HF-2022-P10

17. Welsh Government. A Healthier Wales: our plan for health and Social Care [internet]. 2021 [cited 2025 August 1]. Available from: https://www.gov.wales/sites/default/files/publications/2021-09/a-healthier-wales-our-plan-for-health-and-social-care.pdf

18. Welsh Government. A smoke-free Wales: Our long-term tobacco control strategy [internet]. 2022 [cited 2025 August 1]. Available from: https://www.gov.wales/tobacco-control-strategy-wales-html

19. Public Health Wales. Evaluation of Whole Systems Approach to Healthy Weight [internet]. 2025 [cited 2025 August 1]. Available from: https://phw.nhs.wales/publications/publications1/evaluation-of-whole-systems-approach-to-healthy-weight-report/

20. Welsh Government. Sport and Active lifestyles: state of the national report. National survey for Wales 2022-23 [internet]. 2023 [cited 2025 August 1]. Available from: https://www.sport.wales/files/3be397244ccaa4b4e73dbf613ad98379.pdf

21. Brady M, Brown P. Factors influencing appropriate referrals from NHS 111 to 999 services in Wales. British Journal of Healthcare Management 2024; 30(7)1–12

22. Roynon R, Brady M, Noblett P, Malin, R, Brown C, Fivaz C. Appropriateness of NHS 111 Wales outcomes—using the Call Prioritisation Streaming System: a RAND/UCLA modified Delphi method. BMJ Open 2025;15:e097914.

23. Health Economic Unit. Evaluating the impact and cost benefits of NHS 111 [internet]. 2025 [cited 2025 August 1]. Available from: https://healtheconomicsunit.nhs.uk/case_study/evaluating-the-impact-and-cost-benefits-of-nhs-111/

24. Pope C, Turnbull J, Jones J, Prichard J, Rowsell A, Halford S. Has the NHS 111 urgent care telephone service been a success? Case study and secondary data analysis in England. BMJ Open 2017;7:e014815.

25. Aromataris E, Lockwood C, Porritt K, Pilla B, Jordan Z, editors. JBI Manual for Evidence Synthesis [internet]. JBI; 2024 [cited 2025 August 1]. Available from: https://synthesismanual.jbi.global.

26. World Population Review. High-income countries [internet]. 2025 [cited 2025 August 1]. Available from: https://worldpopulationreview.com/country-rankings/high-income-countries

27. Tricco A, Lillie E, Zarin W, O’Brien K, Colquhoun H, Levac D, et al. PRISMA Extension for Scoping Reviews (PRISMA-ScR): Checklist and Explanation. Ann Intern Med. 2018 Oct 2;169(7):467–473.

28. Pollock D, Peters M, Khalil H, McInerney P, Alexander L, Tricco A, et al. Recommendations for extracting, analysing, and presenting results in scoping reviews. JBI Evid Synth. 2023 Mar 1;21(3):520–532.

